# Molecular pathways associated with Kallikrein 6 overexpression in colorectal cancer

**DOI:** 10.1101/2020.12.09.20245571

**Authors:** Ritu Pandey, Muhan Zhou, Yuliang Chen, Dalila Darmoul, Conner C. Kisiel, Valentine N. Nfonsam, Natalia A. Ignatenko

## Abstract

Colorectal cancer (CRC) remains one of leading causes of cancer-related death worldwide. The high mortality of CRC is related to its ability to metastasize to distant organs. Current management of the CRC disease is limited due to the need of more extensive molecular characterization of tumor biomarkers. Kallikrein 6 (KLK6) is a member of the fifteen-gene family of kallikrein-related peptidases. KLK6 is overexpressed in CRC and contributes to cancer cell invasion through its proteolytic degradation of the extracellular matrix and recently discovered regulation of metastasis signaling pathways.

The goal of this study was to evaluate the clinical features, CRC molecular subtypes, mutational and gene expression patterns of the CRC tumors with overexpressed KLK6 in order to identify KLK6-associated markers for the CRC prognosis and treatment. RNA-Seq data from the CRC patients with a significantly elevated KLK6 transcript levels (KLK6-high group) (Z-score higher than-1.96) were identified in the Cancer Genome Atlas (TCGA) database and processed for clinical, molecular evaluation and bioinformatic analysis, using Gene Ontology (GO), Phenotype and Reactome enrichment and protein interaction methods. KLK6-high cases had a distinct spectrum of mutations in titin (TTN), APC, K-RAS and MUC 16 genes. Differentially expressed genes (DEGs) unique to KLK6-high group were clustered to regulatory pathways controlling the cell signaling, extracellular matrix organization, and cell communication regulation. The members of kallikrein family (KLK7, KLK8, KLK10), keratins (KRT6A, 6B, 15, 16, 19, 80), extracellular matrix proteins (integrin 4B), small proline rich repeat proteins (SPRRs), S100A families, protein trafficking genes (SYL1) and signaling genes within TGF-β, FOS and Ser/Thr protein kinase pathways were recognized as the top KLK6-interaction partners. Expression of selected KLK6-associated genes was validated in a subset of paired normal and tumor CRC patient-derived organoid cultures.

The performed analyses identified KLK6 itself and a set of genes, which are co-expressed with KLK6, as potential clinical biomarkers for the management of the CRC disease.

## Introduction

Colorectal cancer (CRC) manifests from the accumulated defects in oncogenes and tumor suppressor genes, which alter downstream molecular pathways regulating cell proliferation, differentiation, and apoptosis (Wong *et al*., 2007; Fearon, 2011). One of the key hallmarks of cancer is an ability of transformed cells to invade surrounding tissues, which occurs through induction of cell- and tissue-specific signaling pathways [1]. Proteases, which catalyses the hydrolysis of peptide bonds, have long been associated with colon cancer progression because of their ability to degrade extracellular matrices. Tissue kallikrein-related peptidases (KLK) are a subgroup of serine proteases, known to serve a variety of functions, ranging from the normal physiological processes to different disease conditions, including cancer [2]. Several members of kallikrein family are overexpressed in colon cancer patients, i.e. KLK4, KLK6, KLK7, KLK10 and KLK14 [3-6]. Because of association of kallikreins with malignancy they have been proposed as potential diagnostic/prognostic and therapeutic markers [7]. Particularly, KLK6 mRNA levels correlated with serosal invasion, liver metastasis, advanced Duke’s stage, and overall poor patients’ prognosis [8]. Presence of KLK6 transcripts in lymph nodes of CRC patients was associated with the shorter average survival time after surgery and was more sensitive indicator of poor prognosis than carcinoembryonic antigen (CEA) [9]. As we previously reported, KLK6 expression in colon cancer can be induced by oncogenic *K-RAS*, which is the one of the major colon cancer driver genes [9-12].

The aim of our study was to analyze Tissue Cancer Genome Atlas (TCGA) RNA-Seq data from the colorectal cancer samples with high KLK6 expression with the goal to define KLK6-specific gene signature in the CRC for identification of potential markers of tumor progression in colorectal metastatic disease. Our analysis revealed a set of genes, which were differentially expressed in the CRC tumors, which overexpressed KLK6. The set includes genes involved in the regulation of cell signaling, cell-cell communication and proteolysis. The results were confirmed in another GEO dataset (GSE39582). We also analyzed the expression of KLK6 and selected co-expressed genes in the subset of colonic organoids developed from the patients’ surgical tissue (paired normal and tumor cultures).

## Materials and Methods

### RNA-Seq Data Processing

Colorectal Cancer samples from The Cancer Genome atlas (TCGA) were downloaded from Genomic Data Common Website (GDC at https://gdc.cancer.gov/). A total of 480 tumor samples were assayed for transcription. We used the GDC data transfer tool client and GDC API to download all the RNA-seq raw counts data, metadata and available clinical data. Data was analyzed using R (v 3.) and open source methods implemented in R. Raw HTSEQ counts data was normalized using Variance Stabilizing Transformation (VST) method [13] and data was processed further. Z scores were calculated for KLK6 expression across samples and high (N=16) and low (N=7) groups were defined by ≥1.96 and <-1.96 cutoff values, they represent statistically significant outliers. Differential analysis was done using the DESEQ2 [14]. Pearson correlation coefficient was used to identify genes that correlate with KLK6 amongst high and low samples. GEO dataset GSE39582 was downloaded and the array data for 566 colon tumor samples was analyzed for high KLK6 expression. 30 samples were found to be outliers with high KLK6 expression (z score >1.96). Genes results from TCGA high KLK6 tumors were compared to high KLK6 tumors in GSE39582.

### Survival analysis of samples with overexpressed KLK6

The normalized data was combined with clinical data for high KLK6 samples (16), low KLK6 (7) *vs*. other groups (N=457) for TCGA data. The overall survival (OS) in KLK6 high samples compared to other samples was done by utilizing Kaplan-Meier survival plot. GEO dataset was divided into top and bottom quartiles based on KLK6 expression and for high KLK6 samples (30) *vs* others (536). Logrank test was applied to assess the statistical significance between the survival curves. The survdiff function under rms library in R was used to perform the log-rank test.

### Enrichment analysis

Differentially expressed genes (DEG) were analyzed for Gene Ontology using enrichGO from Cluster Profiler and Reactome pathways gene enrichment analysis using enrichPathway from ReactomePA in Bioconductor. Protein interaction data was analyzed using String data resource [15]. The list of differential protein coding genes between high and low KLK6 were searched against Stringdb. The network was filtered for first interacting partners of kallikreins and their associations. Plots: All plots and heatmaps were generated using gplot2 library and other data visualization packages in R.

### CRC patient-derived organoid cultures

The 3D cultures were generated from the fresh surgical material collected as part of the University of Arizona Cancer Center (UACC) Tissue Acquisition and Molecular Analysis Shared Resource (TACMASR) Biorepository efforts. The CRC patients were consented for the tissue collection and all individual patient-related reports and data were de-identified before processing for the tissue biobanking and developing of organoid cultures. Because all information related to the used surgical material was de-identified and did not include any protected health information (PHI), it is not considered a clinical research according on the Human Subjects Protection Program (HSPP) determination. Tissue collection was approved by the Biorepository Oversight Committee at the UACC. The paired normal and tumor specimens, retrieved from the resected surgical material were verified by a certified pathologist. The collected material was placed in the cold Advanced DMEM media and transported to the lab on ice, where they were processed for establishment of organoid cultures as described elsewhere [16]. The organoid cultures generated from paired normal and tumor tissue samples of eight CRC patients were analysed in this study.

### Quantitative Reverse-Transcription Polymerase Chain Reaction

Organoid cultures were washed with AdDMEM/F12 media (Thermo Fisher Scientific. Onc. Carlsbad, CA) once and processed for total RNA isolation using RNeasy® Mini Kit (Qiagen GmbH, Germany, Cat. 74104) according to the manufacturer instructions. Reverse transcription to produce cDNA template was completed using the Applied Biosystems High Capacity cDNA Reverse Transcription Kit (Part #4368814). Quantitative Reverse-Transcription Polymerase Chain reaction (qRT-PCR) was performed using TaqMan® probes (Applied Biosystems, Thermo Fisher Scientific, Inc.) specific for the mRNAs of interest: KLK6 (Hs00160519_ml), KRT6A (Hs00749101_s1), KRT6B (Hs04194231_s1), FOSL1 (Hs04187685_m1), MYC (Hs00153408_m1), HMGA2 (Hs04397751_ml). 0.2 µg of total RNA was reversed transcribed into cDNA in a 20 µL reaction with random hexamers under thermal condition recommended by the protocol. Real-time PCR amplification was performed with the ABI PRISM 7700 SDS instrument (Applied Biosystems, Life Technologies, Inc.), under the universal thermal cycling conditions recommended by the Assay-on-Demand products protocol. Negative controls without template were included in each plate to monitor potential PCR contamination. The expression of genes was tested in triplicate and each reaction was run in duplicate. To determine the relative expression level of each target gene, the comparative *C*_*T*_ method was used. The *C*_*T*_ value of the target gene was normalized by the endogenous reference β2-microglobin (β2M, FAM (Hs99999907_m1)). The relative expression of each target gene was calculated via the equation 2^-Δ*C*^_*T*_ where Δ*C*_*T*_ = *C*_*T*(target)_ – *C*_*T*(endogenous control)_.

### Enzyme-Linked Immunosorbent Assay (ELISA) for KLK6

An ELISA kit for the detection of human KLK6 was obtained from Boster Immunoleader (Pleasanton, CA). The assay was performed according to manufacturer’s protocol. Conditioned media was collected from the cultured organoids on days 2,4,7, and 10 after subculture. KLK6 levels were expressed as picograms per milliliter of media. The plate was read at 490 nm within 30 min of assay ending on a Synergy 2 Multi-Detection Microplate Reader (Bio-Tek Instruments, Inc., Winooski, VT).

## Results

### Pattern of Kallikrein related peptidases family expression in CRC patients

In this study we initially analyzed the relative expression pattern of the members of kallikrein-related peptidase genes family in 480 CRC tumor samples from TCGA. We noted that the transcript levels of a majority of kallikrein-related peptidases vary significantly in the normal and tumor samples amongst the CRC patient’s. **(Fig. 1A**). The normalized transcript levels of KLK6,7,8, 10, 11 and 12 were elevated in tumor samples, compared to normal ones, the levels of KLK2,5,9, and 14 were not altered, and levels of KLK2,3,4, and 13 were found to be lower in tumors, compared to normal tissues. Focusing on KLK6, we further selected the samples that have unusually high or low *KLK*6 expression. We normalized row data using Variance Stabilizing Transformation (VST) method [13] and identified outlier samples with >+-1.96 z score as samples with high and low *KLK*6 expression, respectively. Using these criteria, sixteen samples were identified as high KLK6 expressers and seven samples were selected as low KLK6 expressers. These groups were named as KLK6-high and KLK6-low groups, respectively. Relative expression of other kallikreins family members was plotted for these two groups **(Fig. 1B** upper panel **-** KLK6 -high group, and **Fig. 1B** lower panel **-** KLK6-low group). The above analysis revealed the distinct pattern of expression of KLK family members in KLK6-high group, with particularly high KLK10 co-expression, followed by KLK7, and KLK8. We also noted that KLK10 transcript levels were significantly higher than KLK6, KLK7 and KLK8 in normal tissues (**Fig. 1A**), and KLK10 was elevated in tumor samples in KLK6-low group but to the lesser degree than in KLK6-high samples (**Fig. 1B** lower panel).

**Fig. 1.**
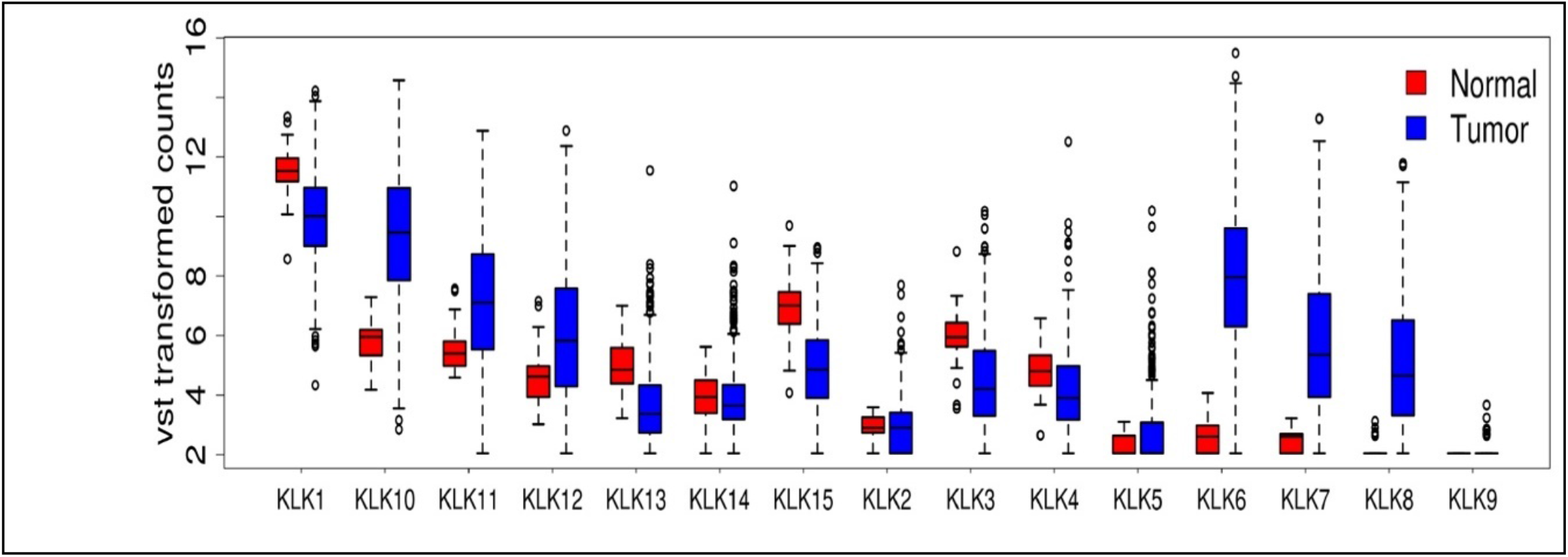
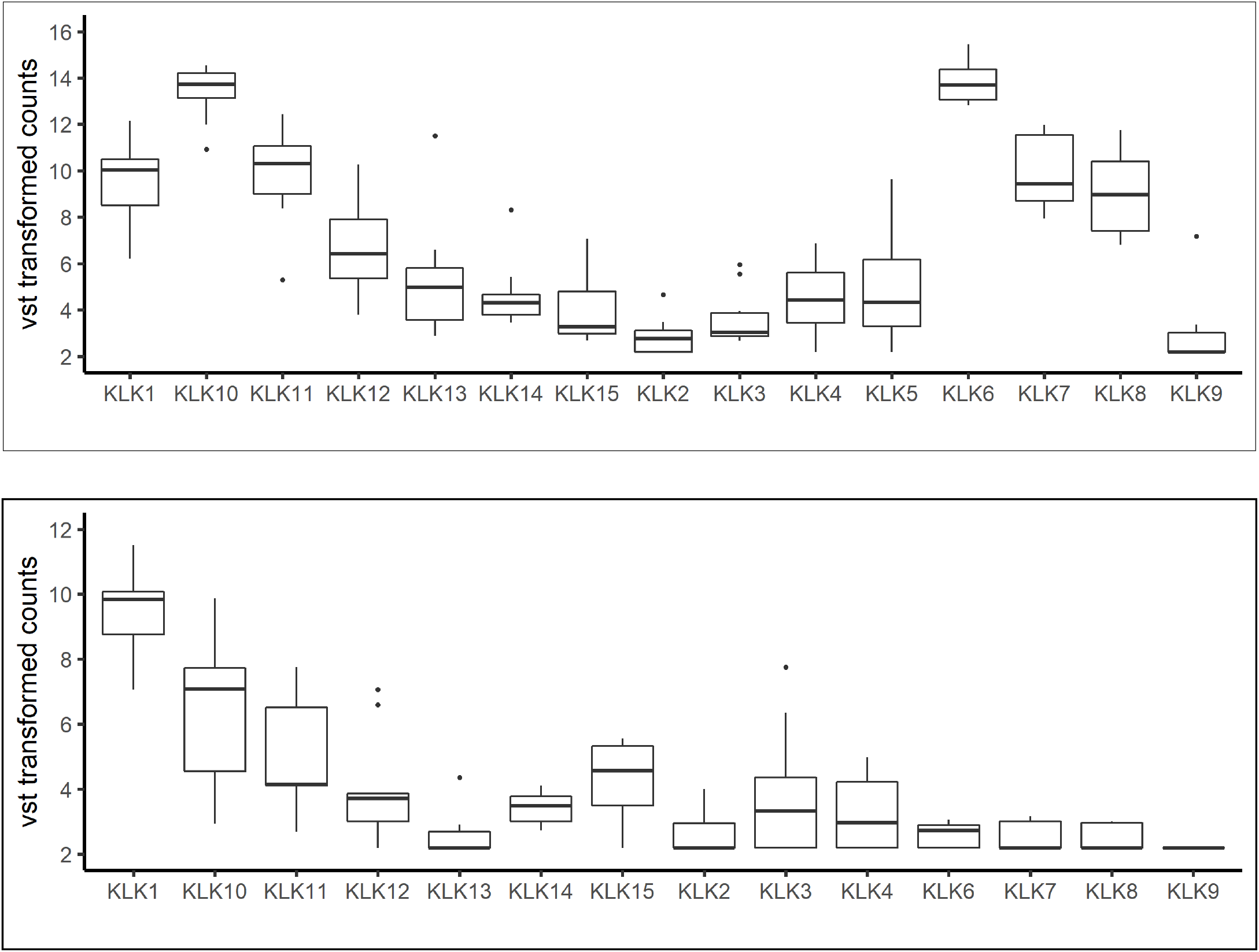
Expression of all members of Kallikrein related peptidase family in colon adenocarcinomas TCGA patient samples. **(A)** Box plot shows expression of KLK gene family normalized by Variance Stabilizing Transformation (VST) method in colon tumors and their relative expression in normal tissues. **(B)** Box plots of KLK family from the CRC patient samples that have been identified as KLK6-high group and KLK6-low group. **Upper panel:** Box plot of expression of all Kallikrein related peptidases in samples with high KLK6 expression (KLK6-high group) and **Lower panel**: Box plot of expression of all Kallikrein related peptidases in samples with low KLK6 expression (KLK6-low group)

### Clinical and molecular characterization of KLK6-high expressing samples

The clinical, gender, histopathological and molecular characteristics for TCGA patients with KLK6-high and KLK6-low (expression are presented in **Table 1)**. The tumors in both groups were identified as colon adenocarcinomas. In KLK6-high group, the number of patients with the advanced stages of the disease constituted more than third of cases (stage III and stage IV: 18.8% and 25%, respectively). In contract, more than half patients in KLK6-low group had the early stages of the disease (stage I and stage II: 28.8% and 43%, respectively), with no stage IV patients.

**Table 1.**
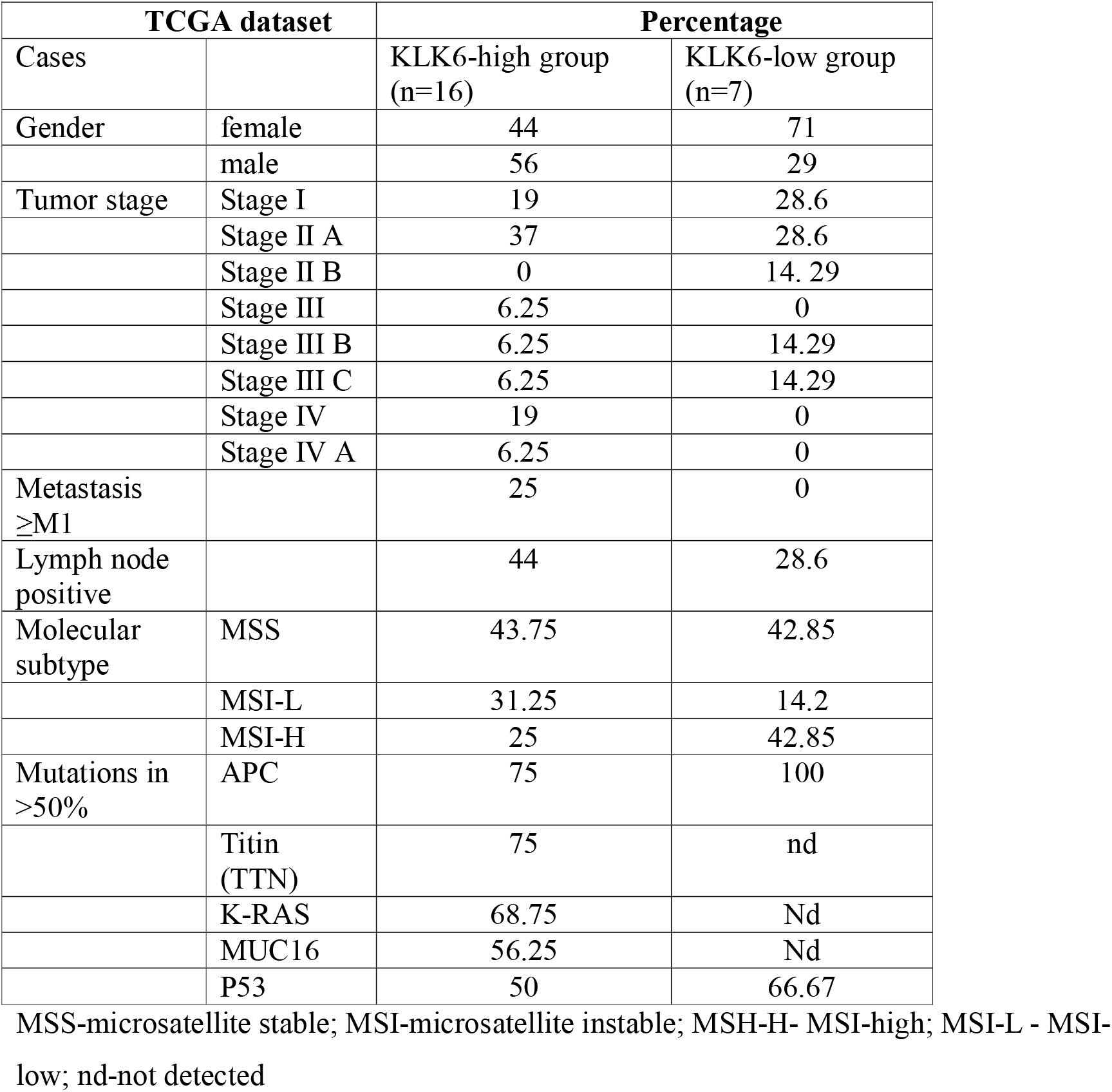
Clinical and molecular analysis of the TCGA colon adenocarcinoma KLK6-high and KLK6-low groups.

### Molecular subtypes in KLK6-high and KLK6-low CRC TCGA samples

The CRC tumors with high and low KLK6 expression were stratified into the microsatellite stable (MSS) and microsatellite instable (MSI) molecular subtypes as defined in other TCGA studies [17]. Both groups had comparable numbers of MSS tumors. Stratification of MSI tumors with high or low number of mutations (MSI-H) and MSI-L, respectively) did not identify any correlation with KLK6 expression (**Table 1**). Similarly, no association or correlation with any particular CMS (Consensus Molecular Subtype) [18] (**S1 Fig.)** was found for high KLK6 expression in tumors.

### Mutation frequencies in KLK6-high and KLK6-low CRC samples

We further determined the spectrum of somatic gene mutations in the KLK6-high and KLK6-low groups by analyzing the available TCGA DNA sequencing results. As demonstrated in **Table 1** and **Fig. 2A**, (based on **S1 Table A** data), the most frequent cancer-related mutations observed in KLK6-high group were mutations in the titin (TTN) and APC genes (found in 75% of KLK6-high tumors), followed by mutations in K-RAS oncogene (68.75%) and MUC16 (mutated in 56.25 %). The mutations in p53 tumor suppressor gene were detected in half samples with high KLK6 expression (**Table 1**).

**Fig. 2.**
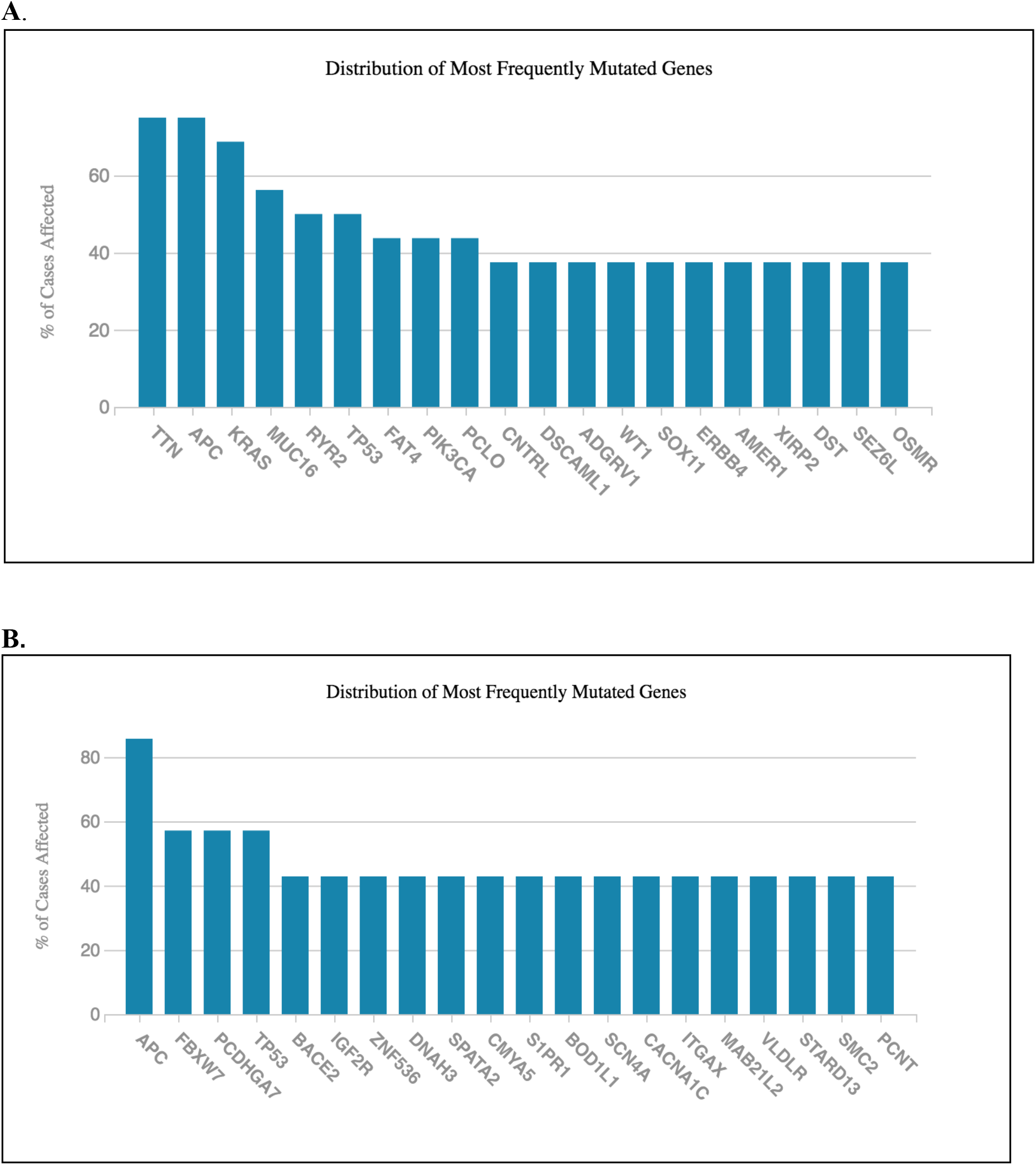
Mutation frequencies in the colon adenocarcinoma TCGA samples with the differential KLK6 transcript level. **(A)** KLK6-high group, **(B)** KLK6-low group.

In contrast, 100% of tumors with low KLK6 expression had APC mutations 66.67% of tumor had p53 mutations. No mutations in TTN or K-RAS were detected in KLK6-low group (**Table 1, Fig. 2B** and **S1 Table B**).

We also assessed the status of KLK6 gene in KLK6-high and KLK6-low samples and did not find any changes in KLK6 gene copy number in these groups. Overall, twenty-five tumors within the remaining 469 cases of the TCGA cohort had alteration in KLK6 gene, with 80% of the cases carrying the missense mutations in KLK6 coding region and 20% of the cases having deletions in the 3’UTR or splice region of KLK6 (**S1 Table C**). This analysis indicates that overexpression of KLK6 in the CRC samples from the TCGA can’t be attributed to alterations in the KLK6 gene copy number or gene’s structural aberrations.

### Survival analysis

Kaplan-Meier survival curves of patients with high KLK6 from TCGA demonstrated that they had overall poor survival compared to other patient samples, although the *p* value did not reach significance **(Fig. 3A)**. Patients with a significantly low KLK6 expression (KLK-low group) had no deaths reported. Because of the limited number of samples with the significantly elevated levels of KLK6 in TCGA, we evaluated the OS rates of the CRC cases from the GEO dataset, GSE39582 of 566 samples. The clinical, histopathological and molecular characteristics of the samples in this dataset are summarized in **S2 Table**. Analysis of GEO samples with the high KLK6 expression levels (30 samples with z score > 1.96) showed a statistically significant decrease in OS rates for these patients (**Fig. 3B**) compared to all other patients (536 samples) (P=0.0066). Dividing the GEO samples into top and bottom quartiles based on KLK6 expression also showed that top quartile has poor survival compared to other groups (P = 0.01) (**Fig. 3C**).

**Fig. 3.**
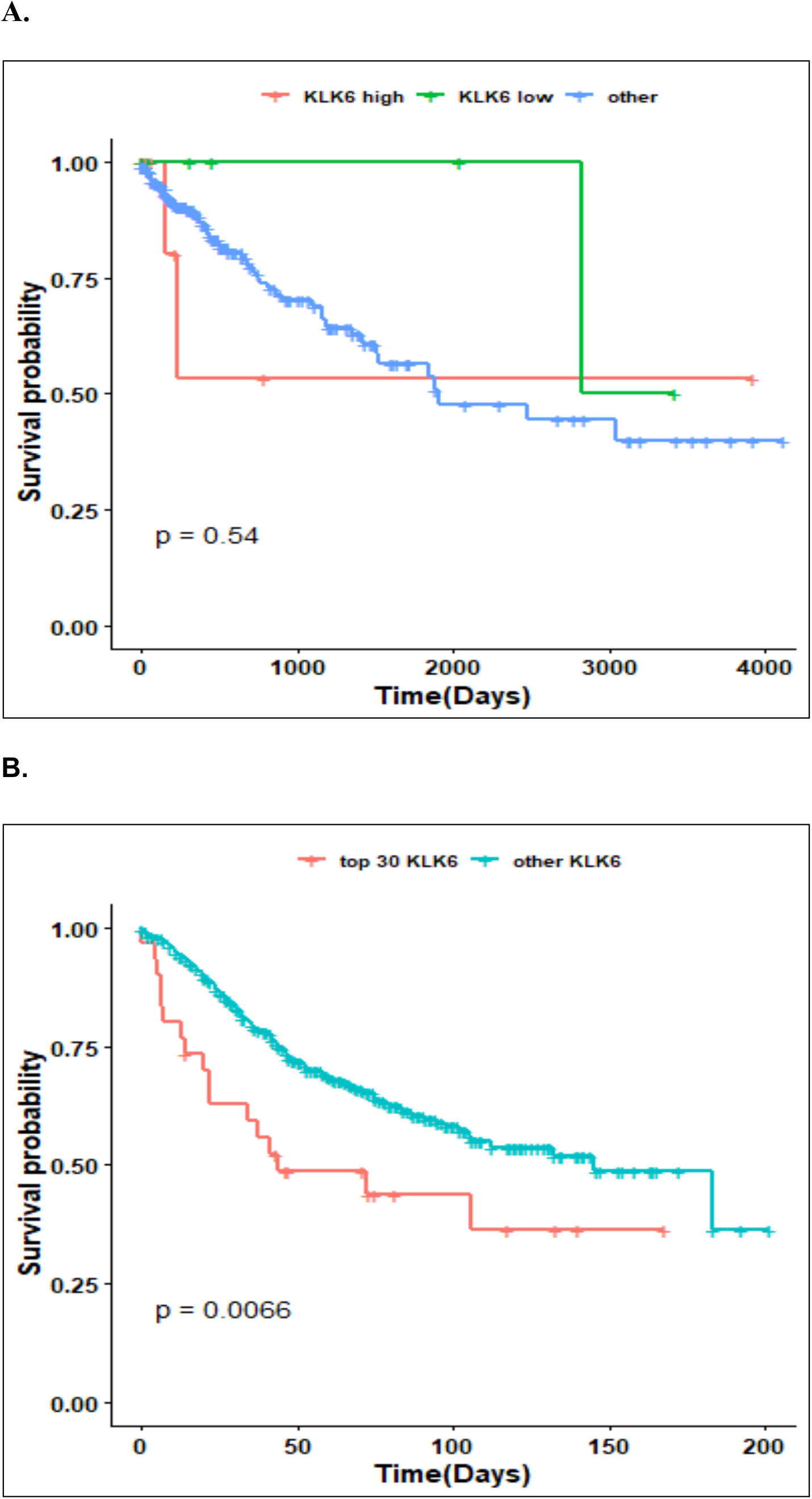
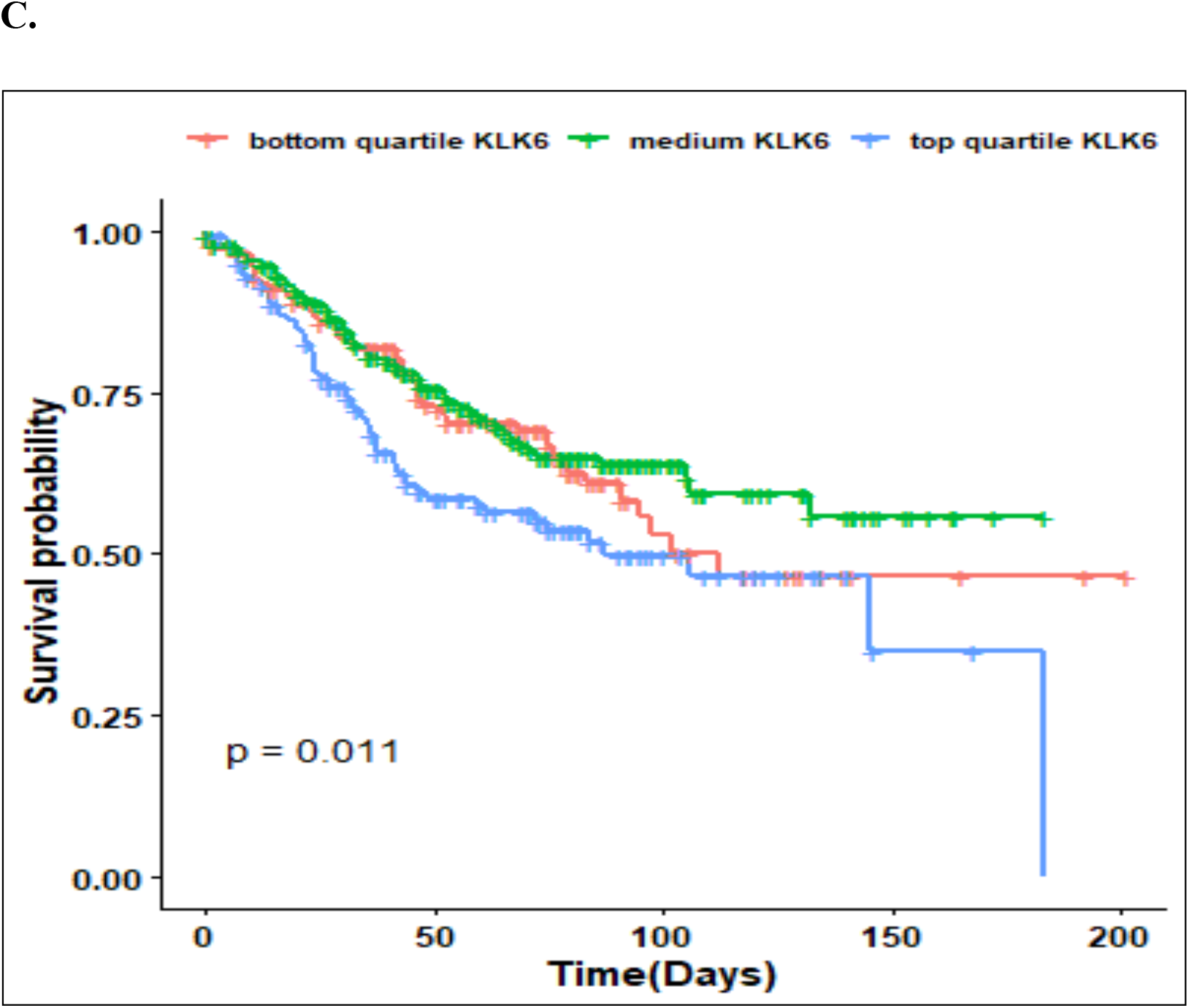
Overall Survival analysis of patients based on KLK6 gene expression from TCGA and GEO datasets. **(A)** Kaplan-Meier Plot of colon adenocarcinoma samples from TCGA. KLK6 High (16 samples), KLK6 low (7 samples) and others were plotted for Overall Survival. (**B)** Kaplan-Meier plot of GEO dataset for KLK6 high samples vs others. (**C)** Kaplan-Meier plot of GEO dataset: based on KLK6 gene expression, data was plotted for upper quartile, bottom quartile and intermediate expression for overall survival.

### KLK6 expression in left-and right sided CRC cases

The right-sided and left-sided colon cancer differ in their clinical and molecular features, and the right-sided colon cases have the worse prognosis [19]. We stratified TCGA CRC cases by the KLK6 expression in the ascending (right) and descending (left) portions of the colon. The KLK6 expression was highly variable within the right, left and transverse colons with no significant difference between tumor locations. The expression of KLK6 was significantly higher in all tumor areas compared to the normal tissues (p=0.001, normal vs descending tumors, p=1.6×10^−6^, normal vs ascending tumors) (**Fig. 4**).

**Fig. 4.**
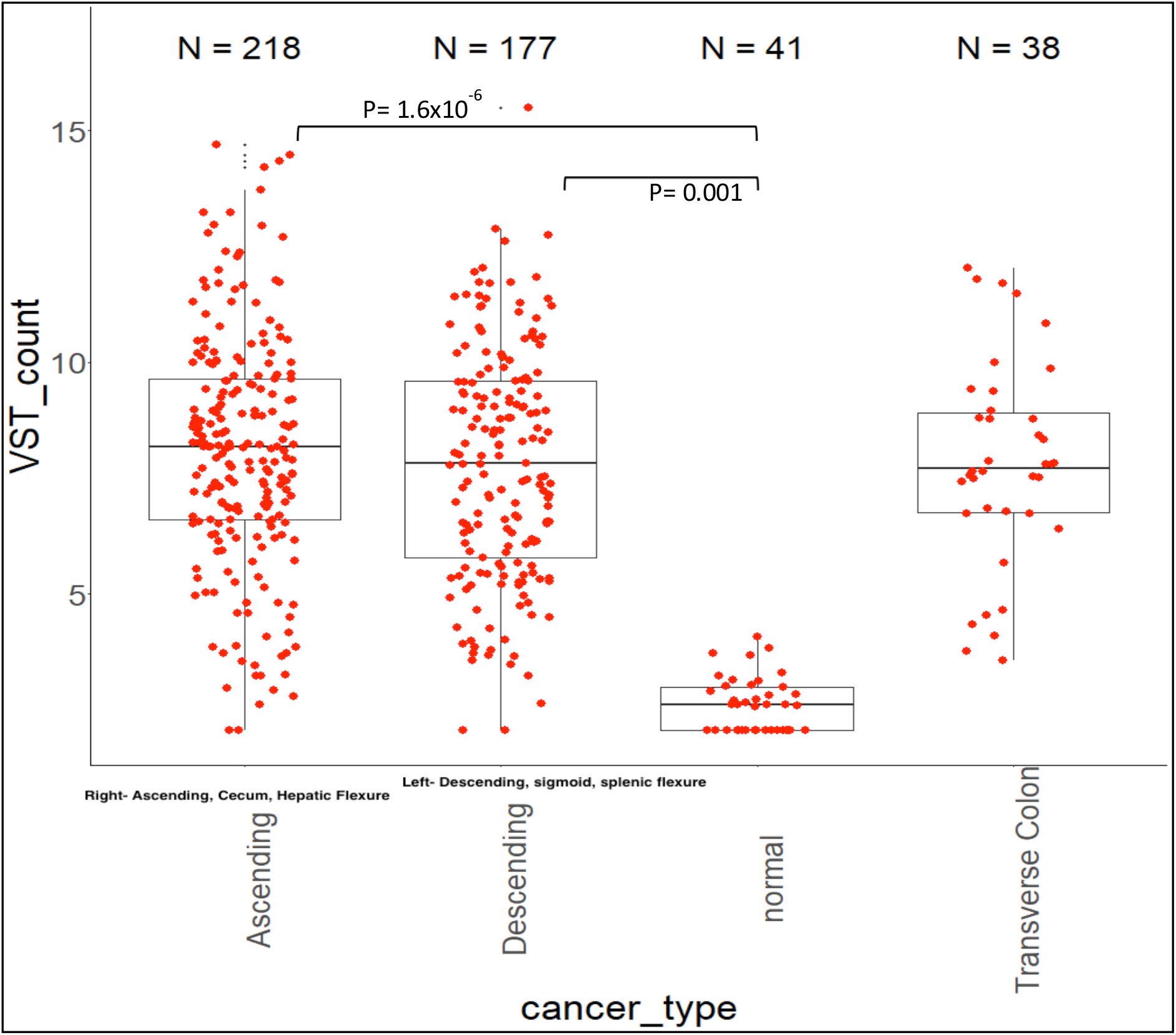
Analysis of KLK6 expression in the right side, left side and transverse CRC cases from TCGA. Box plot shows distribution of KLK6 expression across site of colon adenocarcinoma tumor and normal samples.

### Differential analysis of samples with overexpressed KLK6

A differential expression analysis was done to assess the differences in gene expression between the KLK6-high and KLK6-low patient samples. DESEQ2 analysis revealed 620 differentially expressed genes (DEGs) between these groups (**S3 Table A)**. The DEGs set contained 475 protein coding genes and 145 long non-coding RNAs as well as antisense and uncharacterized transcripts. Cluster analysis of altered gene expression between the KLK6-high and KLK6-low groups showed the KLK6-dependend pattern of gene expression across the groups (**Fig. 5A**). Further differential expression analysis was done between KLK6-high group and the rest of the CRC cases to determine whether any specific genes were altered in KLK6-high group compared to all other samples in the tumor dataset. In the list of all DEGs we identified 236 protein-coding genes which were altered in KLK6-high group (**S3 Table B**). A heatmap of these 236 genes, shown in **Fig. 5B**, demonstrates the pattern of KLK6-dependent gene expression in protein coding genes. Depicted in **Fig. 5C** Venn diagram demonstrates no overlapping in gene expression between KLK6 -high cases and KLK6-low cases, with 171 genes upregulated genes and 54 downregulated genes in KLK6 high group compared to KLK6-low samples and other samples. Therefore, this set of genes is uniquely attributed to the high KLK6 expression in CRC samples. Correlation analysis of KLK6 across high and low KLK6 groups identified a set of transcripts that strongly correlate with KLK6 expression and showed same expression trend (correlation of 0.6 and p value <0.05 were used as significance of association). The correlation plot of top genes, which co-express positively or negatively with KLK6 overexpression is shown in **Fig. 5D**. The highly associated genes include the members of kallikrein family, KLK7, KLK8, KLK10, and KLK11, keratins 80, 6A, 6B, 19, 15, 19, extracellular matrix proteins, vesicle-mediated protein trafficking and with signaling genes, e.g. BMP (TGF-β signaling pathway), GJB3 (Gap Junction protein beta 3), and signaling genes and transcription factors **(S3 Table C**).

**Figure 5.**
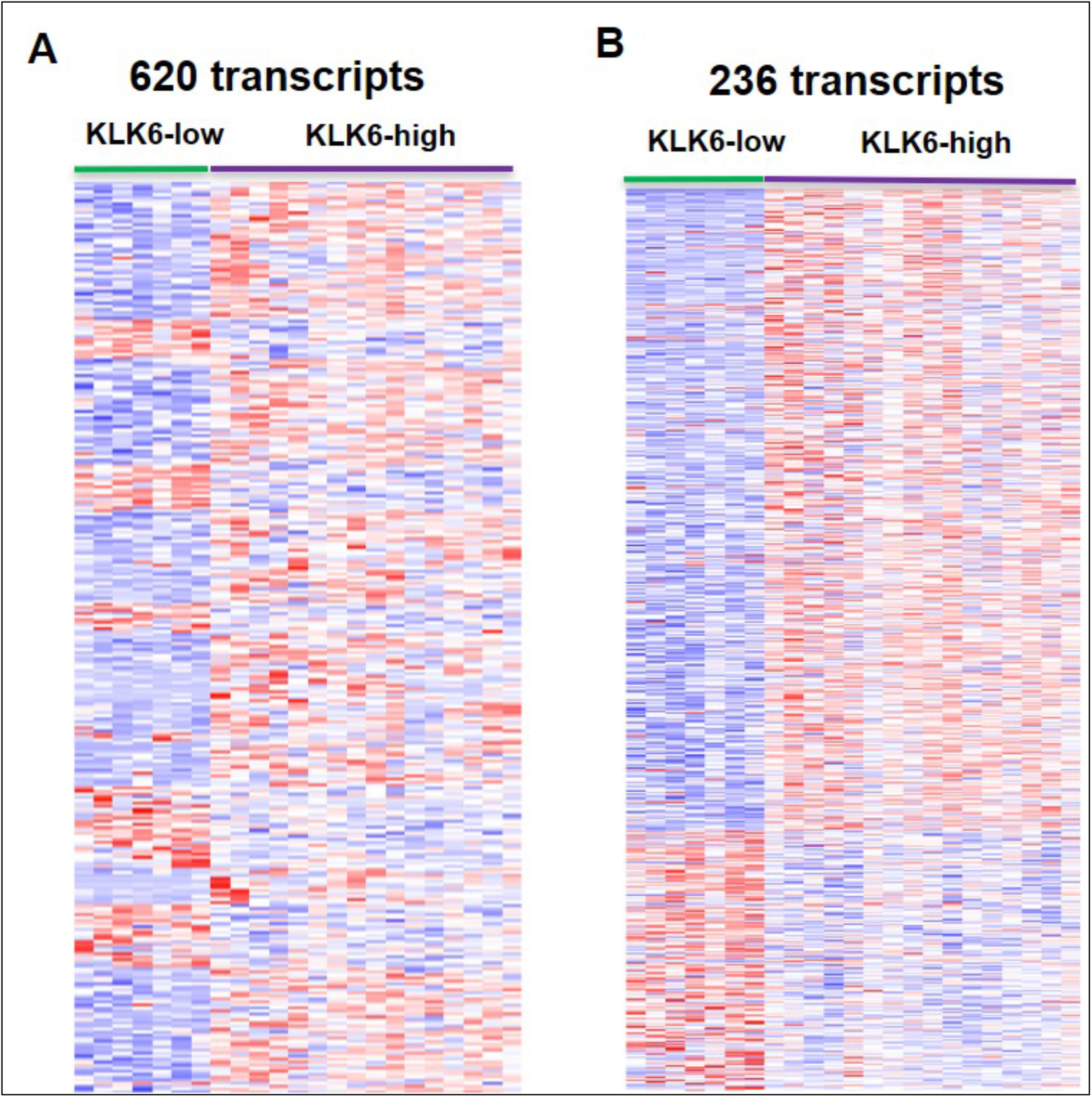
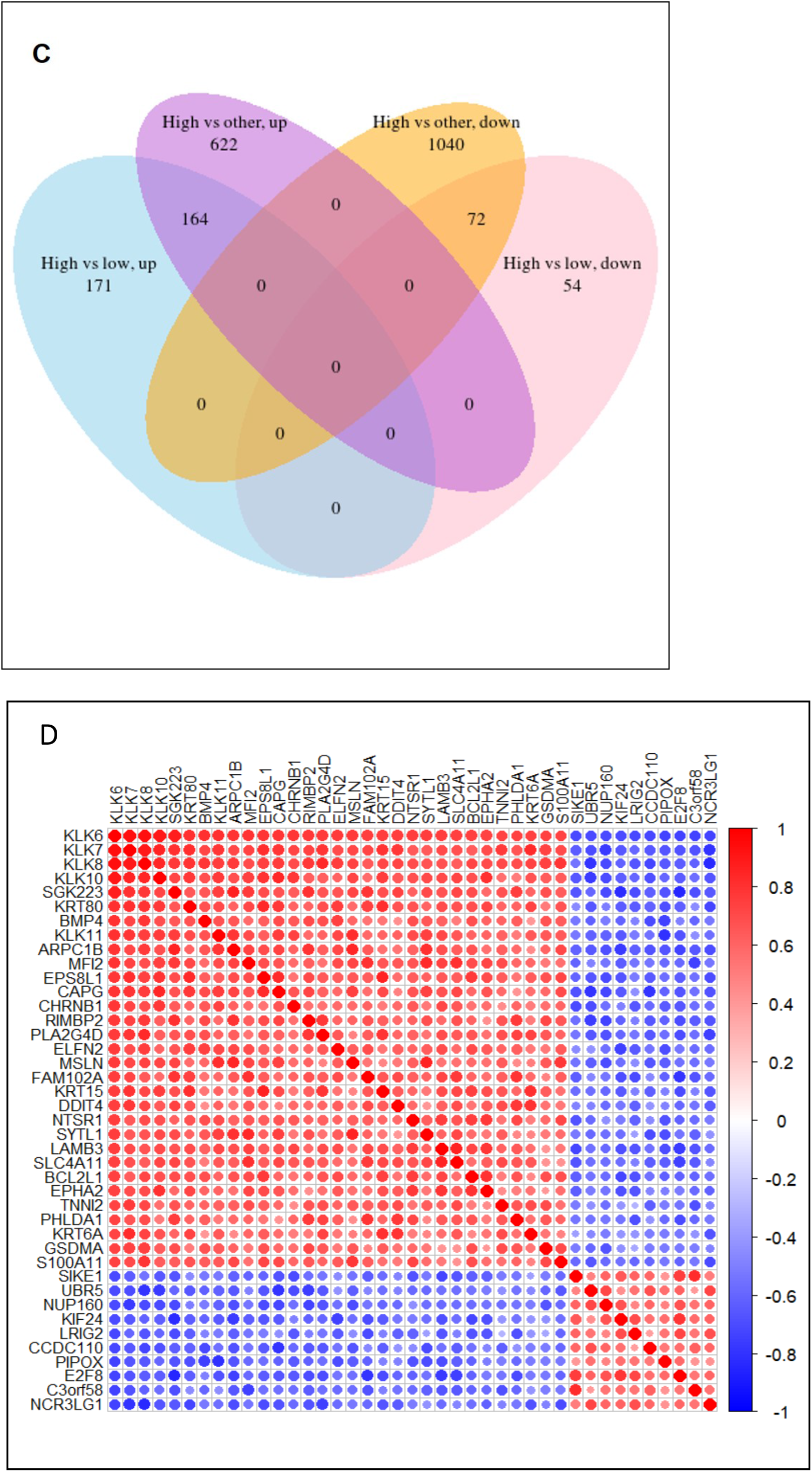
Clustering of the colon adenocarcinoma TCGA samples with differential expression of KLK6. **(A)** Heatmap showing differentially expressed protein-coding genes in KLK6-high vs KLK6-low group samples. **(B)** Heatmap showing a group of 236 differentially expressed protein-coding genes, which are common in high KLK6 vs low KLK6 group and high KLK6 vs other samples. **(C)** Venn diagram showing number of genes which were found to be up and down when KLK6-high group was compared with all other CRC cases. D) Correlation plot of genes co-expressed with KLK6 in KLK6-high group.

Differential expression analysis of high KLK6 samples (30 samples) and other samples (536) in GEO dataset (total of 566 samples) identified 691 significantly altered genes. When we compared the set of 236 KLK6 high specific genes from TCGA dataset with 691 genes from GEO dataset, we found 121 common and the functionally significant genes between the two comparisons **(S3 Table D)**.

### Pathways and significant cellular functions that associated with KLK6 overexpression

We next applied Gene Ontology (GO) and pathway enrichment analyses to find which GO terms are over-represented and under-represented in TCGA KLK6-high DEGs set. In the KLK6-high group we found the most significant association (P value <0.05) of GO terms for biological process (**Fig. 6A**) with the genes involved in response to stimulus, regulation of signaling and regulation of cell communication. **Fig. 6B** demonstrates significantly enriched GO cellular component genes in KLK6-high group involving the plasma membrane, both basolateral and basement, gap junctions and connexon complex as well as membrane-bound vesicle and extracellular exosome. Protein binding genes and peptidase activity (**Fig. 6C**) were significantly enriched for GO molecular function terms **(S4 Table A)**.

**Figure 6.**
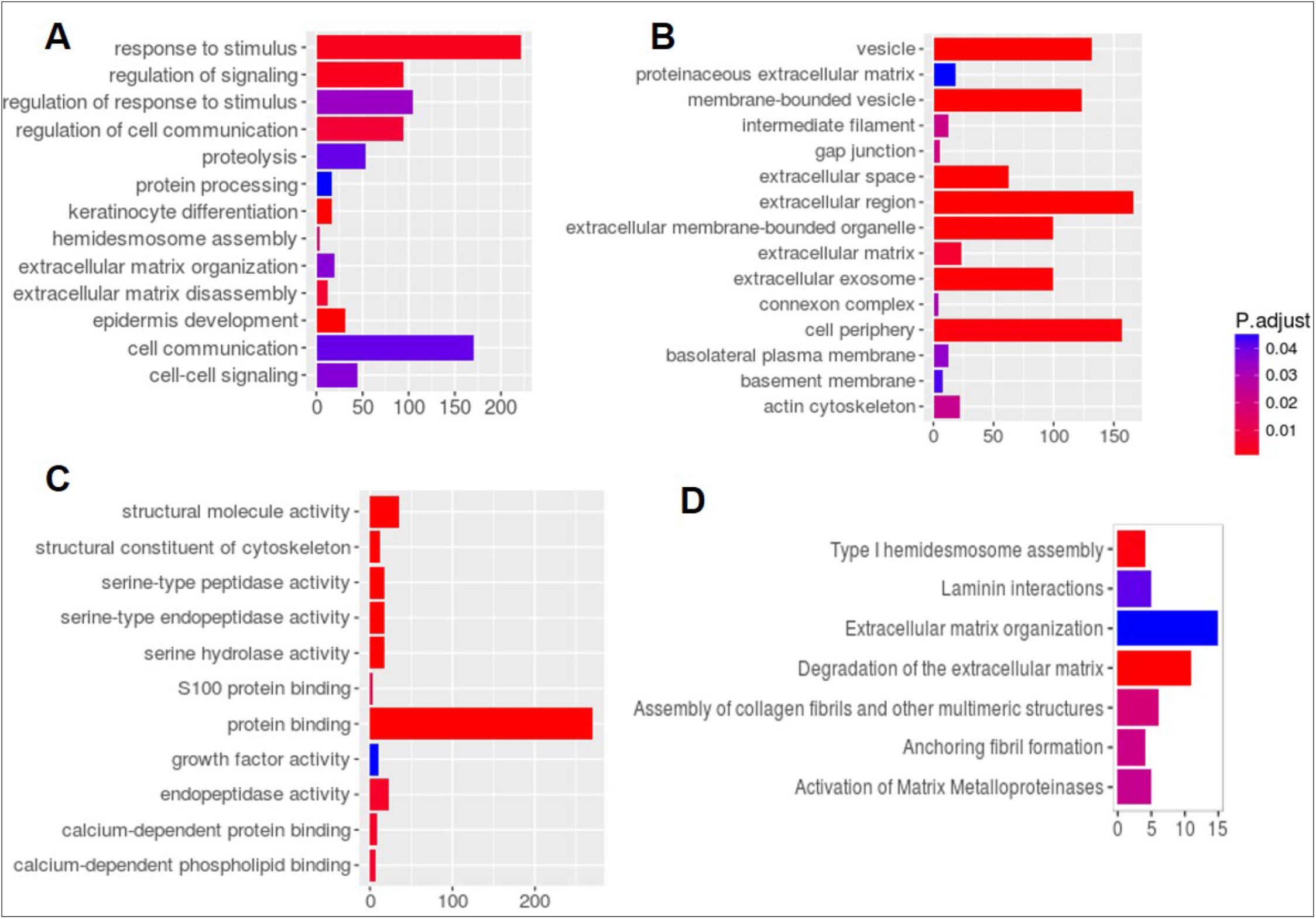
Analysis of Gene Ontology (GO) and signaling pathways associated with KLK6 overexpression in TCGA colon adenocarcinoma samples. Analysis of DEGs for GO shows **(A)** Biological processes. **(B)** Cellular Components and C) Molecular Functional gene sets that are significantly enriched in high expressing KLK6 group. **(D)** Reactome pathways that are enriched in DEGs.

Reactome pathways analysis [20] was applied to search for pathways and several gene sets in Type I hemidesmosome assembly, Laminin interactors, extracellular matrix organization, degradation of the extracellular matrix, assembly of collagen fibrils, Anchoring fibril formation and Activation of Matrix metalloproteinases were found to be differentially enriched in KLK6 high expression samples (**Fig. 6D, S4 Table B**).

Finally, the network of kallikreins interacting genes (KLK6,7,8,10,13) and their first interacting partners was identified using Stringdb **(Fig. 7 and S4 Table C)**. We report here the top 34 extracellular matrix proteins with protein binding function. The list includes metalloproteases, serine proteases, keratins and proteins involved in keratin differentiation (**S4 Table D)**.

**Fig. 7.**
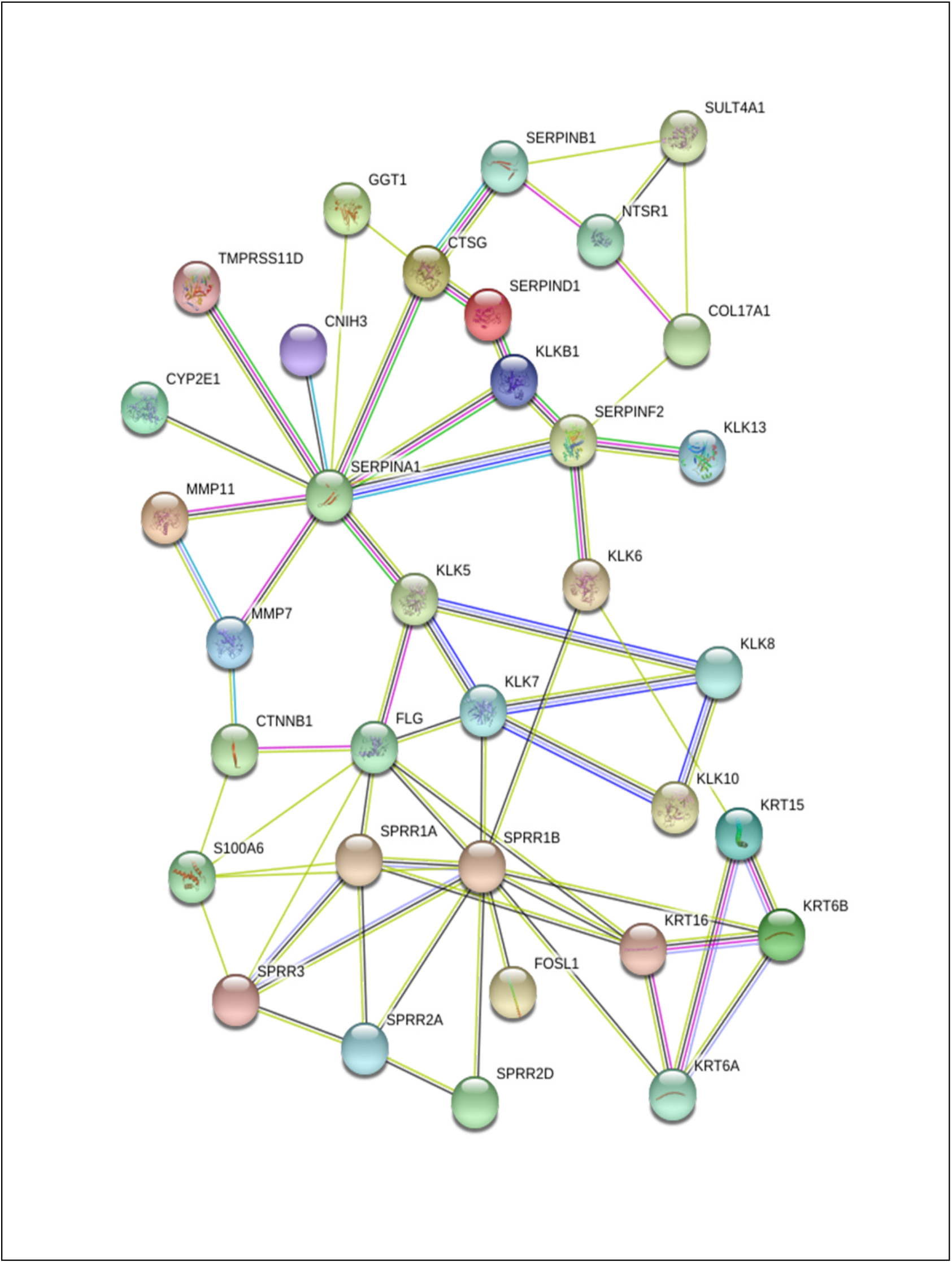
Protein Interaction network of kallikriens and differentially expressed genes found in high KLK6 samples using Stringdb interaction dataset.

### Analysis of expression and secretion of KLK6 in the CRC patient-derived organoid cultures

We analyzed RNA levels of *KLK6* and co-expressers *KRT6A, KRT6B* and *FOSL1* in the paired normal and tumor organoid cultures established from the surgical material of the CRC patients, which were previously untreated. Additionally, because of the importance of *c-MYC* oncogene in colon tumorigenesis [21] and the association of HMGA2 with KLK6 expression and poor disease outcome in the CRC [22, 23], *c-MYC* and *HMGA2* transcripts levels were measured in these culture. Clinicopathological characteristics of the tumors processed for organoid cultures are presented in **S5 Table**. We found the variable levels of *KLK6* transcript in the screened cases with no association between KLK6 expression and the molecular subtype of the analyzed tumors (**Fig. 8A**). The *HMGA2* and *FOSL1* transcript levels were detected in 4 out of 7 cases and the *FOSL1* level was found to be low compared to others genes. The *KRT6A* and *KRT6B* gene transcripts were significantly elevated in the adenoma (one case) compared to malignant tumors (**Fig. 8B**). The *c-MYC* transcript level was significantly higher in P#2T and P#13T organoid cultures (both MSS subtype and stage T3N0) along with overexpression of *KLK6, HMGA2*, and *FOSL1*, which suggests the aggressive phenotype of these tumors. We also measured the levels of secreted KLK6 in the conditioned media of organoids derived from the normal and tumor tissue samples using KLK6 ELISA assay (**Fig. 8C**). The analysis of the secreted KLK6 levels in three cases, representing the colon cancer samples (P#3 and P#5) and a colon adenoma (P#7) showed that the elevated levels of secreted KLK6 were detected in the tumor cultures compared to normal ones and they increased with the time in culture. The secreted KLK6 was below the level of detection in the normal cultures of P#3 and P#7. The detected secreted KLK6 in the normal culture of P#5 suggests that the tissue has acquired some invasive properties. Overall, this pilot analysis confirms the variability of KLK6 expression in the CRC and demonstrates the experimental platform for more detailed analysis of key genes in the colon cancer.

**Figure 8.**
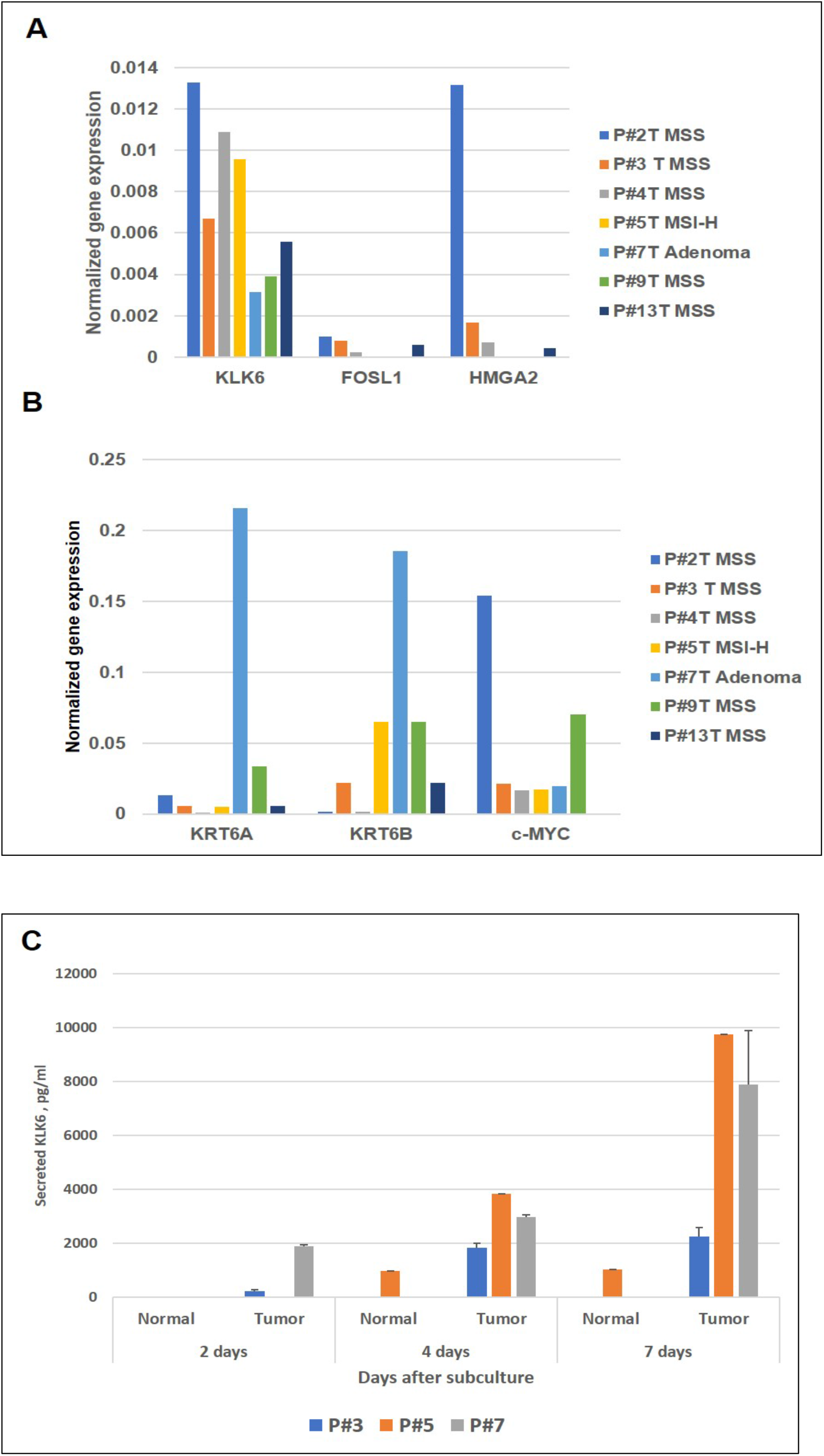
Evaluation of the transcript levels of selected co-expressed genes and KLK6 secretion in the CRC patient-derived organoid cultures. **(A)** of KLK6, FOSL 1 and HMGA2 transcript levels by quantitative RT PCR. **(B)** Expression of keratin 6A and 6B and c-Myc. **(C)** Levels of secreted KLK6 in the conditioned media of the tumor organoid cultures by ELISA.

## Discussion

The ongoing challenge in cancer treatment is to identify the most appropriate targeted therapy for managing the disease based on the individual properties of the tumor. This precision medicine approach requires identification of patients individual genetic, epigenetic and other alterations for further stratification into subgroups with matching drug treatment options. Kallikreins have become a subject of intense investigation as prognostic markers and candidates for drug targeting because of their important functions in normal physiology and pathological state, which have been discovered in the past years (reviewed in [24]). Recent analysis of kallikrein gene family across 15 different cancers was done with a goal to identify the suitable biomarkers within kallikrein gene family and highlighted KLK6, KLK7, KLK8 and KLK10 as the excellent diagnostic biomarkers candidates for colon adenocarcinoma [25]. Another analysis of TCGA dataset, which focused on search of prognostic biomarkers in colon adenocarcinomas, identified KLK6 gene as one of the two overexpressed genes (MYC was the second one), which are significantly associated with overall survival in adenocarcinoma patients and therefore have high accuracy in predicting the disease outcome [26].

In our study, we selected the extremely high KLK6 expressing cases (KLK6-high group, 16 cases) from TCGA database and analyzed the clinical parameters and molecular pathways and genes, which are associated with KLK6 overexpression. KLK6-high CRC cases in TCGA revealed that 25% of patients in this group had an advanced clinical stage (Stage IV and IVA), while none patients in KLK6-low group were diagnosed with Stage IV of the disease. No difference in the molecular subtypes of the adenocarcinomas were noted between KLK6-high and KLK6-low cases. At the same time, the analysis of the mutations revealed that the KLK6-high cases had the high frequencies of mutations in titin (TTN), APC, K-RAS and MUC 16 genes, while in KLK6-low group APC mutations were found at 100% frequency and TP53 mutation was identified in 66% of cases. We have previously reported the mutant K-RAS-dependent expression of KLK6 gene in the colon cancer cells [11, 27], so the current results from TCGA patient data confirm the validity of the previous findings.

The mutational landscape in the KLK6-high group suggests the possibility of utilizing the tumor mutational burden (TMB) biomarker for estimating the therapy outcomes in the CRC patients with high KLK6 expression. The TMB is a novel and a promising biomarker used for predicting responsiveness to Immune Checkpoint Blockage (ICB) [28, 29]. Tumors with high TMB, independent from MSI-H status, such as KLK6 high expression tumors, are not well studied. Titin (TTN), a structural protein in striated muscles [30] and MUC16, which encodes the cancer antigen CA-125, are the two longest genes in the genome. The correlation between the high TMB and the mutated status of the genes having the larger size has been previously suggested [31]. Recent analysis of a discovery cohort of 34 solid tumors and 7 cohorts from clinical trials treated with programmed cell-death ligand 1 (PD-L1) inhibitor demonstrated that TTN mutations in solid tumors can be used to stratify into groups with different clinical responses to ICB monotherapy [31]The unique presence of both of these mutations along with high number of K-RAS and APC mutations in the KLK6-high cases from TCGA dataset warrants further validation of the TBM applicability in the CRC patients with KLK6-high cases for estimating their eligibility for ICB therapy in prospective clinical trials. PIK3CA, another one of the most commonly mutated genes in CRC is also found in ∼43% of high KLK6 tumors. In the course of DEGs analysis, a set of the 236 protein-coding DEGs were identified, which is unique to KLK6-high group. The correlation analysis identified the members of kallikrein family, KLK7, KLK8, KLK10, as the most highly co-expressed with KLK6. This finding is in agreement with limited analysis of kallikrein family in the CRC reported by Tailor and co-workers [25]. Co-expression of KLK6 and KLK10 has also been reported in pancreatic adenocarcinomas, where it was defined as an unfavorable factor associated with a poor OS [32].

The other overexpressed genes, which correlated with KLK6, were. keratins 6A, 6B, 15, 16, 19, 80. This set of upregulated keratins is different from the one, identified in KLK6 overexpressing serous ovarian cancer [33]. This finding suggests that KLK6 regulates expression of keratins in cancer in a tissue-specific manner. The list of altered genes included extracellular matrix proteins ING4B (integrin 4B), LAMB3 (laminin subunit beta3), vesicle-mediated transport protein coding genes SYL1 (Synaptotagmin Like 1) and GJB3 (Gap Junction protein beta 3), as well as signaling genes, such as BMP4 (Bone Morphologic Protein 4, in TGFββ signaling pathway) and transcription factor FOSL1 (FOS-like transcription factor subunit). Analysis of the functional roles of the most altered genes showed that they are important structural components of the cell cytoskeleton, gap junction and connexon complexes, the basolateral membrane and actin cytoskeleton, as well as extracellular exosomes, which indicates that KLK6 overexpression contributes to aggressive growth and invasion in tumor cells.

Protein interaction network for KLK6-overexpressing cases allowed us to identify proteins which may interact with KLK6, either through formation of protein complexes or via modifications of other proteins. Particularly, the presence of the kallikrein family members, KLK5, KLK7, KLK8, KLK10, KLK13, and the metalloproteinase family members MMP7 and MMP11 in this network suggests that active KLK6 enzyme may induce activation and cross activation of other proteases. The constructed network predicts the direct interactions between KLK7, KLK8 and KLK10, but not KLK6. This observation highlights complex relations among kallikreins in the CRC. Through protein interaction network we found keratins 6A, 6B, 15 and 16, as well as Small Proline Rich Repeat (SPRR) genes (SPRR1A, SPRR1B, SPRR2A, SPRR2D and SPRR3), which are involved in the keratinization pathway, as interacting partners of KLK6. This suggest the mechanism of KLK6-mediated upregulation of a subgroup of keratin genes in KLK6-high group. Keratins are established epithelial cell markers and can regulate multiple signaling pathways in cancer. The keratins altered in KLK6-high colorectal tumors may be further evaluated as biomarkers in the CRC progression. SPRRs proteins are the keratinocytes differentiation markers, involved in formation of the cornified envelope in skin cells, but their function in cancer is not well defined. The SPRR3 has been reported as tumor suppressor in esophageal squamous carcinoma cells [34] The S100A6 gene, which was found to be an interacting partner of SPRR3 and SPRR1A in KLK6 high group, has been associated with cells motility and tumor metastasis in cervical cancer cells and lung carcinoma [35, 36]. SERPIN F2, a serine protease inhibitor, which was identified in connection with KLK6 overexpression, is known inhibitor of plasmin, trypsin and chemotrypsin

Although in this study we focused on protein-coding genes, we noted top two highly up regulated non-coding transcripts in KLK6 high group samples, i.e. CTB-147C22.8 and CTB147C22, which were reported to be co-expressed with kallikrein genes KLK5, KLK6 and KLK7 in HPV16^+^ head and neck tumors [37]. Using a set of 7 patient derived tumor organoid cultures we confirmed the variability in expression and secretion of KLK6, and necessity of individualized approach in designing the CRC chemotherapy.

The findings presented here suggest that KLK6-high tumors may be sensitive to the specific inhibitors of kallikreins. The reversible inhibitors of the members of kallikrein family have been previously reported [38-40]. Applicable to our targets, the promising inhibitors of KLK6 [41, 42] and KLK7 [43] have been recently characterized. The pre-clinical testing of selected kallikrein inhibitors should be done to validate their use as investigational drugs in clinical trials especially because of demonstrated here high complexity of interactions between kallikreins.

The analysis of KLK6 -overexpressing cases from TCGA was verified using a GEO CRC sample set and the overall findings were in agreement with the results of TCGA analysis. There were higher number of KLK6-overexpression cases in this set (30 samples vs 16 in TCGA) and the GEO patients had more advance stages of the disease (no patients with Stage 1 and the higher percent of patients had Stage II and Stage III of the disease). Perhaps because of these differences, the analysis of OS rates in the GEO dataset showed that patient’s survival rates significantly correlated with KLK6 overexpression in tumors, while in TCGA samples the Kaplan-Meier curves demonstrated a similar trend.

## Conclusions

Overall, applying different types of analysis we discovered the distinct sets of genes, such as the members of kallikreins, keratins, extracellular matrix proteins, SPRRs, S100A families, protein trafficking and signaling genes within TGF-β, FOS and Ser/Thr protein kinase families. These genes are involved in regulation and coordination of colon cancer cell invasion and metastasis in KLK6-overexpressed CRCs. Our analysis suggests that identifying colon cancer patients with KLK6 overexpression and targeting it using specific inhibitors can be beneficial for the suppression of metastasis. In a future we hope to match the altered pathways and genes in the individual patient cases with available or investigational drug treatment options.

## Data Availability

All data generated and analyzed during this study are included in the current article and the supplementary information files.

## List of abbreviations

KLK6: Kallikrein 6
CRC: colorectal cancer
GDC: Genomics Data Commons
GO: Gene Ontology
TCGA: The Cancer Genome Atlas
GEO: Gene Expression Omnibus
KEGG: Kyoto Encyclopedia of Genes and Genomes
DEGs: differentially expressed genes
VST: Variance Stabilizing Transformation
OS: Overall Survival

## Acknowledgements

We would like to thank Dr David B. Stewart, MD, FACS, FASCRS, Professor of Surgery at University of Arizona for facilitating the clinical samples acquisition and Ms. Carole Kepler, B.S., MT (ASCP), Biospecimen Operations Manager of the University of Arizona Cancer for her efforts in the tissue collection program. We also thank the patients who were consent for tissue collection for organoid cultures.

## Authors’ contributions

RP and NI designed the study, interpreted data and wrote the manuscript. ZM and CY performed the bioinformatics data acquisition and analysis and assisted with manuscript preparation. DD was involved in the analysis and interpretation of the data. CK gave technical assistance with *in vitro* experiments. VN supervised the clinical samples acquisition and evaluation. RP and NI are the corresponding authors. All authors read and approved the final manuscript.

## Supporting information

**S1 Figure. KLK6 expression in normal colon and tumors from TCGA stratified according to Consensus Molecular subtypes (CMS)**. CMS1-4 are four subtypes defined as in [18]. NoLBL are TCGA tumors with no labels or could not be subtype classified.

**S1 Table. Supplemental data for molecular characterization of TCGA samples. Data are from Genomics Data Commons (GDC). A)** Top 100 frequently mutated genes in Colon adenocarcinoma TCGA KLK6 high samples. **B)** Top 100 mutated genes in Colon adenocarcinoma TCGA low KLK6 samples. **C)** Analysis of alterations in KLK6 gene in TCGA cohort.

**S2 Table. Patient Characteristics of high KLK6 samples from GEO dataset GSE39582**.

**S3 Table. Differential expression analysis of genes in the KLK6-high and KLK6-low patient samples. A)** Differential Expressed Genes between high KLK6 expressed Group and low KLK6 expressed groups in TCGA Colon Adenocarcinoma samples; **B)** 236 Differential Expressed Genes between high KLK6 expressed Group and low KLK6 expressed groups and high KLK6 expressed groups and rest all the samples in TCGA colon tumor; **C)** Genes correlated with KLK6 in Colon adenocarcinoma TCGA samples. **D)** Genes found in common between TCGA DEG list of 236 genes and GEO dataset GSE39582 of KLK6 high samples compared with rest of the samples.

**S4 Table. Supplemental data for pathways associated with KLK6 overexpression in colorectal cancer. A)** Gene Ontology Terms enriched in KLK6 high samples in TCGA for **Fig. 6C. B)** Reactome Pathways enriched in KLK6 high samples in TCGA for **Fig. 6D. C)** Protein Interaction of Kallikreins and differential expression genes in KLK6 High Samples in TCGA from Stringdb for **Fig.7. D)** Annotation**-**Protein Interaction partners with Kallikreins in **Fig. 7**.

**S5 Table. Clinicopathological characteristics of surgical cases established as organoid cultures**.

