## Supplementary material for "Molecular pathways associated with Kallikrein 6 overexpression in colorectal cancer": S1 Figure

**S1 Figure. KLK6 expression in normal colon and tumors from TCGA stratified according to Consensus Molecular subtypes (CMS).** CMS1-4 are four subtypes defined as in [18]. NoLBL are TCGA tumors with no labels or could not be subtype classified.

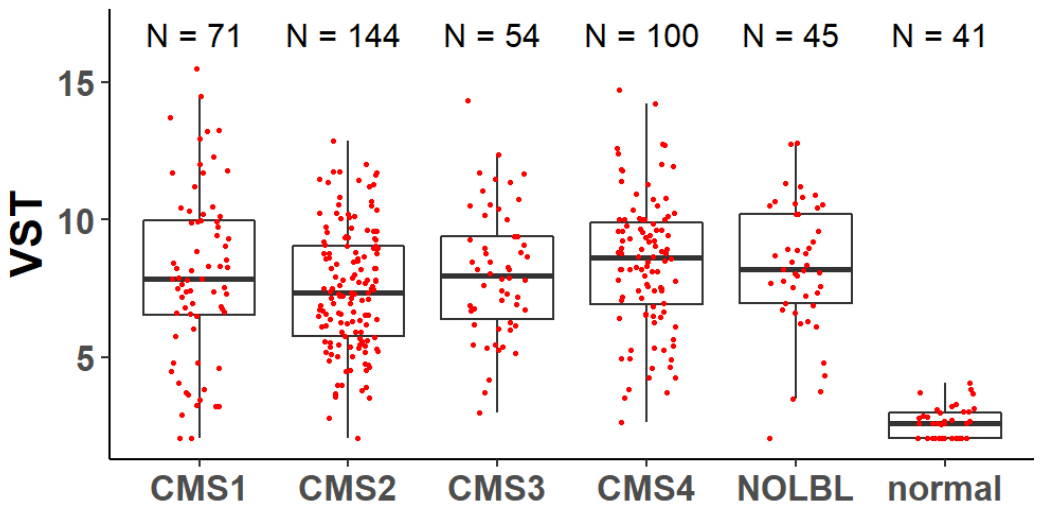
