## Supplementary material for "Molecular pathways associated with Kallikrein 6 overexpression in colorectal cancer": S2 Table

**S2 Table. Patient Characteristics of high KLK6 samples from GEO dataset GSE39582.**

| **GEO dataset GSE39582** | | **Percentage** |
| --- | --- | --- |
| Cases |  | KLK6-high group (n=30) |
| Gender | female | 40 |
|  | male | 60 |
| Tumor stage | Stage I | 0 |
|  | Stage II | 53.3 |
|  | Stage II A/ B | NA |
|  | Stage III | 29 |
|  | Stage III B | NA |
|  | Stage III C | NA |
|  | Stage IV | 16.7 |
|  | Stage IV A | NA |
| Metastasis ≥M1 |  | 16.6 |
| Lymph node positive |  | NA |
| Molecular subtype | MSS | 73.3 |
|  | MSI-L/H | NA |
|  | MSI | 16.67 |
| Mutations in >50% | APC | NA |
|  | Titin (TTN) | NA |
|  | K-RAS | 53.3 |
|  | MUC16 | NA |
|  | P53 | 52.3 |

MSS-microsatellite stable; MSI-microsatellite instable; MSH-H- MSI-high; MSI-L - MSI-low
