## Supplementary material for "Molecular pathways associated with Kallikrein 6 overexpression in colorectal cancer": S5 Table

**S5 Table.** **Clinicopathological characteristics of surgical cases established as organoid cultures*.**

| **De-Identified**  **ID** | **Sex** | **Molecular subtype** | **Clinical info** |
| --- | --- | --- | --- |
| P2 | F | MSS | Sigmoid cancer, G2-G3 moderate to poor differentiated, pT3, pN0 |
| P3 | F | MSS | Colon and rectal adenocarcinoma, G2 moderately differentiated, pT3, pN0 |
| P4 | M | MSS | Rectal adenocarcinoma, G2 moderately differentiated, ypT3N1 |
| P5 | M | MSI-H | Transverse colon adenocarcinoma, G2 moderately differentiated, mpT3 pT2 pNo |
| P7 | M | N/A | Ascending colon polyp, Villous adenoma, not malignant |
| P9 | M | MSS | Left colon adenocarcinoma, G2 moderately differentiated, pT2N0 |
| P11 | F | MSI-H | Right colon adenocarcinoma, tumor, G1 well differentiated, mpT3 pN0 |
| P12 | F | MSS | Colon and rectal adenocarcinoma, G2 moderately differentiated, pT3 pN1b |
| P13 | F | MSS | Right colon adenocarcinoma, G2 moderately differentiated, ypT3N0 |

*All individual patient-related information used for this Table was de-identified.
